# Simulation of trajectories in the illness-death model for chronic diseases: discrete event simulation, Doob-Gillespie algorithm and coverage of Wald confidence intervals

**DOI:** 10.1101/2020.04.01.20049809

**Authors:** Ralph Brinks, Annika Hoyer

## Abstract

We compare two approaches for simulating events in the illness-death model in a test example about type 2 diabetes in Germany. The first approach is a discrete event simulation, where relevant events, i.e., onset of disease and death, are simulated for each subject individually. The second approach is the Doob-Gillespie algorithm, which simulates the number of people in each state of the illness-death model at each point in time. The algorithms are compared in terms of bias, variance and speed. Based on the results of the comparison in the test example, we assess coverage of the corresponding Wald confidence intervals.

## Introduction

For the past decades the world has been experiencing a pandemia of chronic diseases [1-3]. The World Health Organisation WHO has estimated that 71.3% percent of the 56 million worldwide deaths in 2016 were due to non-communicable (chronic) diseases (NCDs). For comparison, in 2000 this percentage has been 60.5% [4]. This has led the United Nations’ General Assembly to set forth the target of reducing the burden of chronic diseases by increase physical activity, reducing salt-intake, and lowering tobacco and alcohol consumption [5]. Possible ways of projecting the impact of such and related health policies usually comprise involvement of mathematical models [6].

Mathematical models about disease development and progression are often expressed as multi-state models. The famous SIR model of Kermack & McKendrick consists of the states *Susceptible, Infected* and *Recovered* (which led to the acronym *SIR*) and is a simple but valuable and frequently used example [7]. For chronic conditions, the illness-death model also consists of three states: *Healthy* (with respect to the condition under consideration), *Ill* and *Dead* [8]. In case there is no remission, the illness-death model of chronic diseases looks like depicted in Figure 1. The transition rates between the states are the incidence rate (*i*), the mortality rates with (*m*_1_) and without the disease (*m*_0_). The rates *i, m*_0_, and *m*_1_ may depend on a time scale τ, e.g. calendar time or the age in a birth cohort.

**Figure 1:**
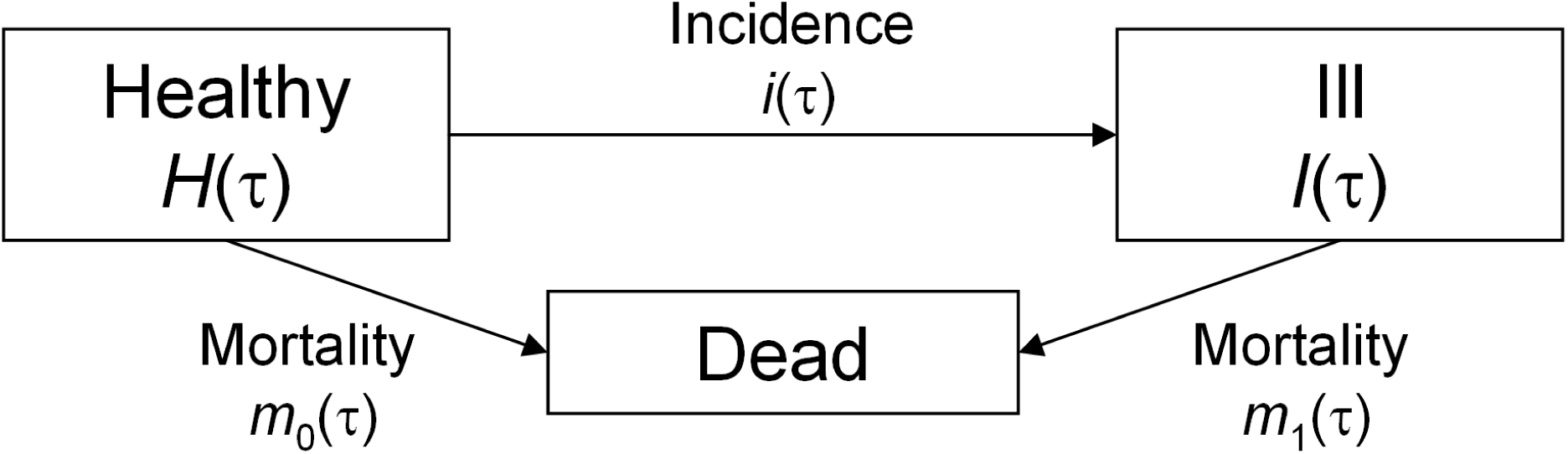
Illness-death model for chronic diseases without remission.

For a given point in time τ, let *H*(τ) and *I*(τ) denote the (absolute) number of people in the *Healthy* and *Ill* state, respectively. Then, we have shown recently that the percentage *p*(τ) = *I*(τ)/[*H*(τ)+*I*(τ)] of people alive in the *Ill* state at time τ is governed by an ordinary differential equation (ODE) [9]:

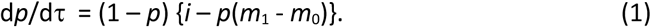

The percentage *p* is also known as prevalence. Equation (1) follows from the Kolmogorow forward equations for time-inhomogeneous Markov processes [10]. Analytical properties of the ODE (1) are studied in [11].

For a single subject, the point in time when a transition from one state in Figure 1 into another takes place, is stochastic. However, by its nature, the ODE (1) is deterministic. The question in which way deterministic models are the limiting cases of stochastic models is more than 50 years old [12]. For certain types of ODEs it could be shown that these ODEs are limits of counting processes [13]. However, ODE (1) is a quotient of counting processes (in *H* and *H+I*) and limit theorems for quotients of random variables (“ratio distributions”) are difficult to treat [14,15]. Thus, we chose a more pragmatic approach to investigate how large a group of individuals must be such that the ODE (1) describes the empirical prevalence *p* “appropriately”. To answer this question, we simulate groups of individuals transiting through the stages of the illness-death model shown in Figure 1.

Two research questions are addressed: First, we compare the commonly used discrete event simulation [16] with the Doob-Gillespie algorithm [17, 18] in terms of bias, variance and computational speed. The comparison is accomplished in a test example motivated from diabetes in the German population [11]. Second, we use the better of the two algorithms to explore the coverage probability of the 95% Wald confidence intervals of the binomial distribution for different population sizes and success probabilities. The success probabilities correspond to the prevalences of the chronic disease in the illness-death model. We chose the Wald confidence interval, because it is easy to calculate and “has acquired a nearly universal acceptance in practice” [19].

## Methods

To decide if the discrete event simulation is faster than the Doob-Gillespie algorithm, we set up a test example. The following transition rates *i, m*_0_, and *m*_1_ are chosen for the illness-death model:

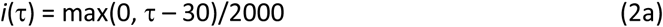

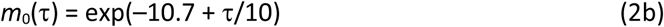

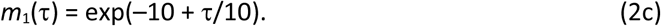

These rates are motivated from the epidemiology of type 2 diabetes in Germany [11, 20]. In order to study groups of individuals transiting through the stages of the illness-death model in Figure 1, two algorithms are compared. The first method is the discrete event simulation, where for each subject the sojourn times in the states *Healthy* and eventually *Ill* are drawn from an appropriate distribution [16]. The second method is the Doob-Gillespie algorithm, where for each (small) time step, the number of transitions between the states are drawn from a Poisson distribution [17,18]. A total of *N* = 500 simulation runs of populations with initial size *n*_0_ = 1000 are computed with discrete event simulation and the Doob-Gillespie algorithm. The resulting bias, variance and computing times are compared.

In a second analysis, the faster of the two methods is chosen to explore the coverage probability of the 95% Wald confidence intervals of the binomial distribution for different population sizes (*n*) and success probabilities (*p*). For this, we use initial population sizes *n*_0_ = 50, 500, 5000, 50000, and 500000. These populations transit through the stages of the illness-death model until a maximal age (here 86 years) is reached. Then, we mimic cross-sectional studies at different points in time τ_k_ = 35, 40, 45, 50, 60, 70, 75, 80, 85 (years). The success probabilities *p* of the binomial distribution correspond to the prevalences *p* of the chronic disease in the cross-sections at times τ_k_. Note that people decease by entering the absorbing state *Dead* in Figure 1, such that the number *n* of people still alive at τ_k_ differs from the initial population size *n*_0_. To estimate coverage, *N* = 10000 simulation runs for each *n*_0_ are accomplished. 95% Wald confidence intervals are calculated by

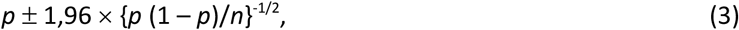

where *p* = *p*(τ) is the solution of the ODE (1) at time τ. The prevalences *p* are obtained from solving ODE (1) with rates (2) and initial condition *p*(30) = 0. Then, the Wald 95% confidence intervals are calculated by Eq (3) at the points τ_k_ = 35, 40, 45, 50, 60, 70, 75, 80, 85. The proportion of trajectories calculated by the Doob-Gillespie algorithm that are included in the Wald confidence interval at τ_k_ is the coverage. Figure 2 shows the flow diagram of the simulation.

**Figure 2:**
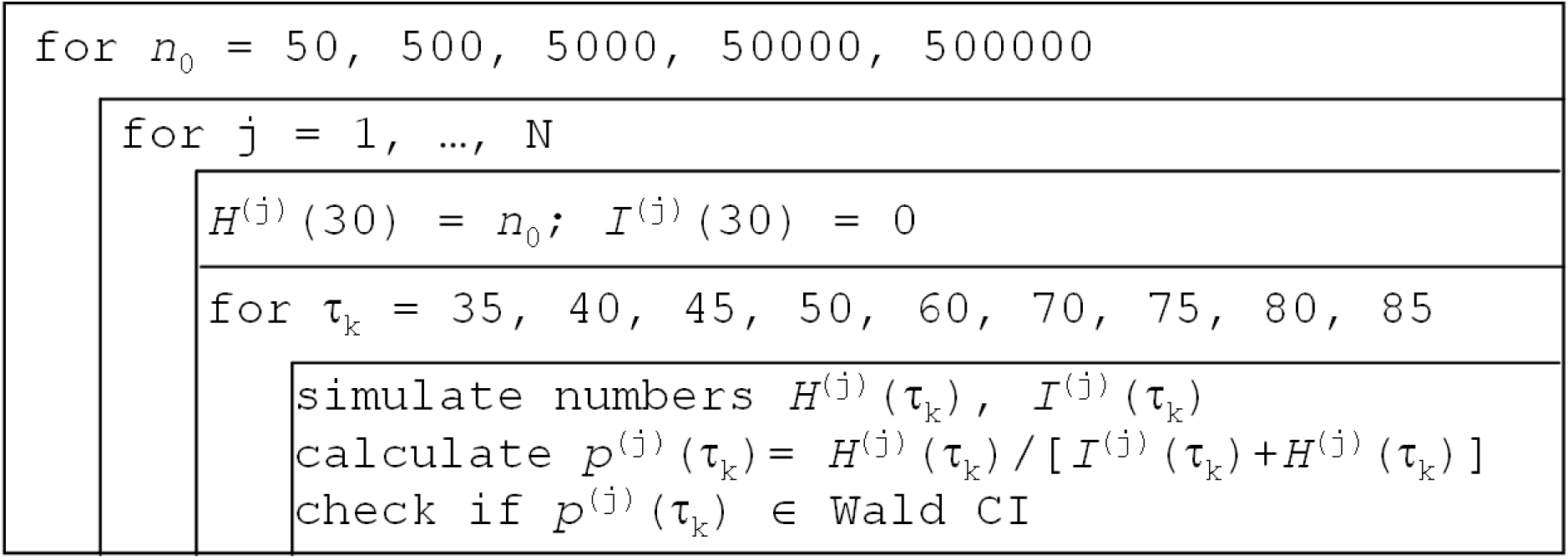
Flow diagram of the simulation.

## Results

Figure 3 shows the slope field for the ODE (1) with rates as in Eq. (2) [11]. In addition, we plotted the solution of the ODE with the initial condition *p*(30) = 0 as a red line.

**Figure 3:**
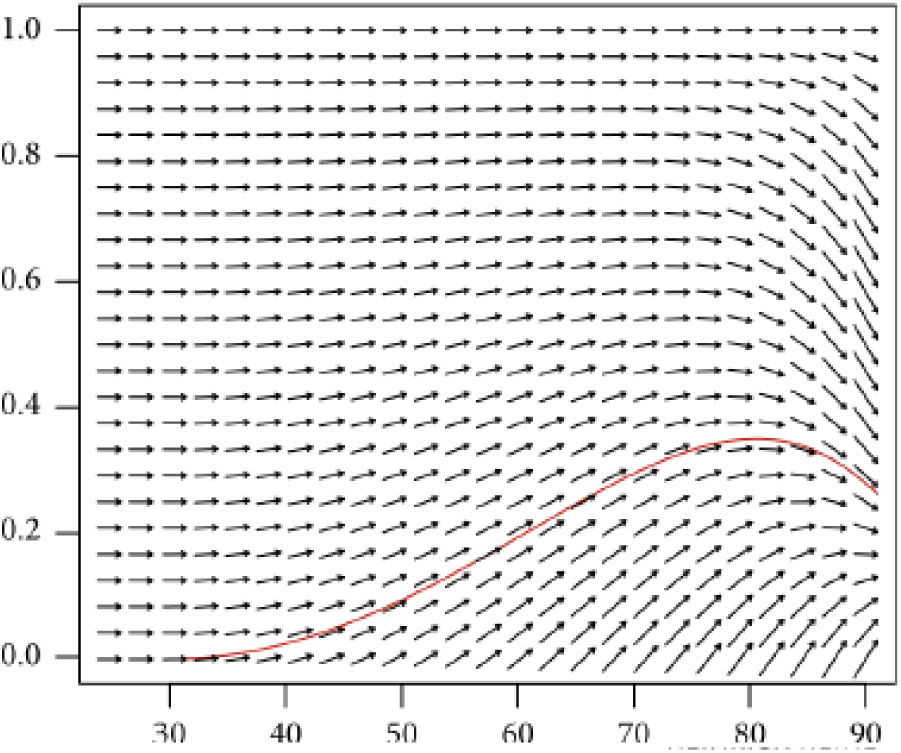
Slope field of the ODE (1) with transition rates *i, m*_0_, and *m*_1_ as in Equations (2). The red line is the solution of the ODE with the initial condition *p*(30) = 0 [Brinks Hindawi].

Figure 4 shows the results of *N* = 500 simulation runs with initial population sizes *n*_0_ = 1000. Each black line corresponds to one of the *N* = 500 simulations of *p*(τ) versus τ. We call these lines prevalence trajectories *p*^(j)^(τ), *j* = 1, …, *N*. The left and right panel of Figure 4 has been computed by the discrete event simulation and the Doob-Gillespie algorithm, respectively. In both panels, all simulated prevalence trajectories start at τ = 30 with *p*(30) = 0. As τ increases, the prevalence increases until about τ = 80 and decreases thereafter. It can be seen that the width of the black area increases as τ increases (“inverted saxophone”). Computing times on a standard personal computer (Intel i3-3220 with 3.3 GHz and 8 GB RAM) have been 65.4 and 0.932 seconds for discrete event simulation and Doob-Gillespie algorithm, respectively. Hence, the Doob-Gillespie algorithm is 70x faster than the discrete event simulation.

**Figure 4:**
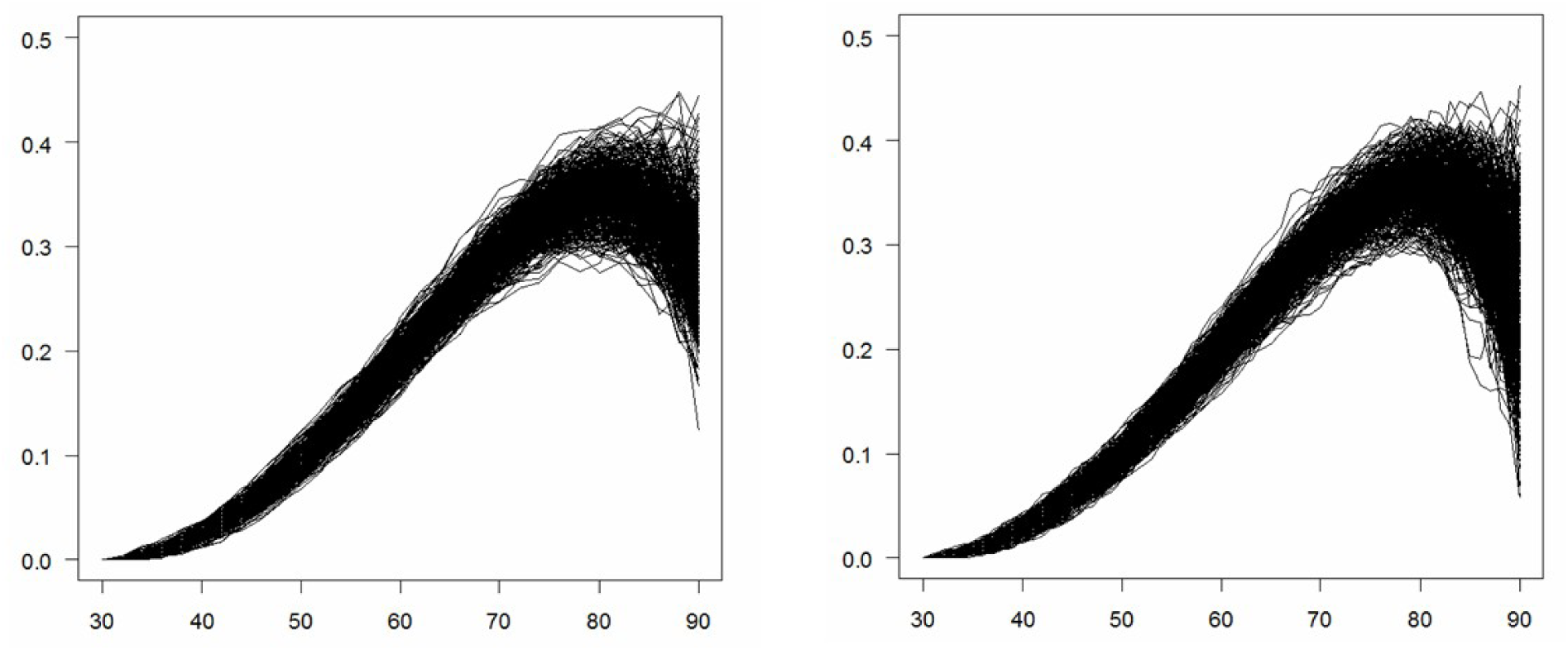
Trajectories of *N* = 500 simulations with discrete event simulation (left) and the Doob-Gillespie algorithm (right).

Figure 5 shows the box plots of the differences between the true prevalence *p* (obtained from solving ODE (1) with rates (2) and initial condition *p*(30) = 0) and the trajectories *p*^(j)^, *j* = 1, …, *N*, at different ages τ_k_. The left and right part of Figure 5 refers to the discrete event simulation and the Doob-Gillespie algorithm, respectively. The empirical mean of the difference (solid lines in the boxes) is close to zero in all ages and both simulations algorithms. Variance of the differences increases as age increases. Thus, in terms of bias and variance, both algorithms yield very similar results.

**Figure 5:**
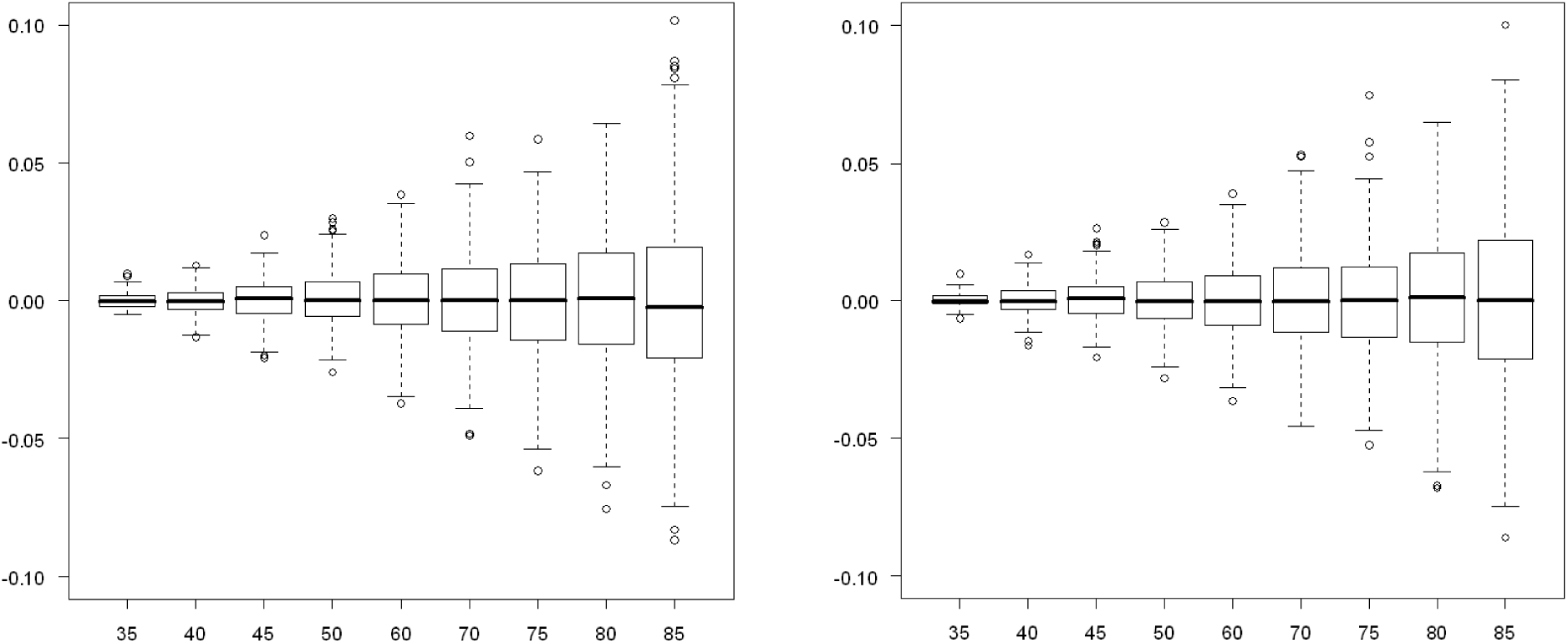
Boxplots of the difference between *N* = 500 trajectories and the true prevalences *p* at different ages τ_k_ (abscissa). The panels show the differences for the discrete event simulation (left) and the Doob-Gillespie algorithm (right).

After the Doob-Gillespie algorithm outperformed the discrete event simulation in terms of computational speed while both algorithms are similar in bias and variance, we continued with the Doob-Gillespie algorithm to examine coverage of the 95% Wald confidence intervals. The results are shown in Table 1. The first row shows the points in time τ_k_, where the coverage is assessed. The second row presents the true prevalence *p*(τ_k_) calculated by solving ODE (1) with rates (2) and initial condition *p*(30) = 0. The third row in Table 1 shows the proportion of people who are still alive at time τ_k_ (*S* means survival function). For instance, at τ_k_ = 75 we find that *S*(τ_k_) = 60.3%, which means that about 40% of the initial *n*_0_ people have died. Thus, the number *n* of people alive at τ_k_ can be calculated by *n* = *n*_0_ × *S*.

**Table 1:** Coverage of the 95% Wald confidence interval as in Eq. (3) in *N* = 10,000 simulations of the Doob-Gillespie algorithm.

| | | $\tau = 35$ | $\tau = 40$ | $\tau = 45$ | $\tau = 50$ | $\tau = 60$ | $\tau = 70$ | $\tau = 75$ | $\tau = 80$ | $\tau = 85$ |
| --- | --- | --- | --- | --- | --- | --- | --- | --- | --- | --- |
| $p(\tau)$ in % | | 0.62 | 2.46 | 5.4 | 9.4 | 19.3 | 29.5 | 33.3 | 35.0 | 33.6 |
| $S(\tau)$ in % | | 99.7 | 99.2 | 98.4 | 97.0 | 90.8 | 74.6 | 60.3 | 42.2 | 23.4 |
| $n_0$ | 50 | 0.9622 | 0.9658 | 0.9455 | 0.9571 | 0.9512 | 0.9499 | 0.9531 | 0.9457 | 0.9463 |
|  | 500 | 0.9617 | 0.9452 | 0.9518 | 0.9519 | 0.9493 | 0.9500 | 0.9486 | 0.9514 | 0.9456 |
|  | 5000 | 0.9449 | 0.9509 | 0.9494 | 0.9516 | 0.9484 | 0.9467 | 0.9461 | 0.9461 | 0.9549 |
|  | 50000 | 0.9529 | 0.9510 | 0.9530 | 0.9555 | 0.9517 | 0.9506 | 0.9500 | 0.9513 | 0.9507 |
|  | 500000 | 0.9476 | 0.9490 | 0.9487 | 0.9492 | 0.9497 | 0.9483 | 0.9491 | 0.9508 | 0.9473 |

Irrespective of the tested simulation settings, the coverage probability of the 95% Wald confidence intervals is at least 94.49% and reaches up to 96.58%. Thus, in the tested settings the 95% Wald confidence interval calculated by Eq. (3) with *p* being the solution of the ODE (1) is a practical and reasonable approximation to the 95% confidence interval.

## Conclusion

Simulations about a test example shows that the 95% Wald confidence bounds calculated by Eq (3) using the ODE (1) for calculating the prevalence *p* have a satisfactory coverage -irrespective of the tested population sizes *n* and the tested magnitudes of *p*. In this sense, the ODE and the associated Wald confidence bounds describes populations appropriately on a wide variety of epidemiological scales.

## Data Availability

This is a simulation study. Scripts for use with the free statistical software R is available from the authors.

